# Changing Epidemiology of Acute Myocardial Infarction in the High-Sensitivity Cardiac Troponin Era

**DOI:** 10.64898/2026.08.26.26361490

**Authors:** Brandon Taylor, Connor Oltman, Jurgen Shtembari, Naveed Adoni

## Abstract

Contemporary national-scale electronic health record (EHR) trends in documented acute myocardial infarction (AMI) rates during the high-sensitivity cardiac troponin (hs-cTn) and Type 2 myocardial infarction (T2MI) era are not well characterized. We conducted a serial cross-sectional analysis of U.S. adults aged ≥18 years in Epic Cosmos from 2016-2024, encompassing 821,859,867 patient-years. Age- and sex-standardized AMI diagnosis rates increased 75.7%, from 343.1 to 602.7 per 100,000 patients. This increase was predominantly driven by T2MI, which increased 133.8% from 99.9 per 100,000 in 2018 to 233.4 per 100,000 in 2024; NSTEMI increased 13.8% while STEMI decreased 4.1%. Annual hs-cTn-tested encounters increased 34.5-fold from 2017 through 2024. The proportion of tested encounters associated with any AMI remained relatively stable after 2021, whereas T2MI continued to increase and surpassed NSTEMI in 2024 as the most frequently diagnosed AMI subtype per hs-cTn-tested encounters. Males had higher absolute AMI rates across all age groups, although relative increases were greater among females. Documented AMI epidemiology shifted substantially toward T2MI during expanding hs-cTn utilization, underscoring the need for evidence-based approaches to the evaluation and management of T2MI.

## Introduction

Acute myocardial infarction (AMI) is a leading cause of morbidity and mortality in the United States, with an estimated 805,000 events occurring annually (1). The epidemiologic landscape of AMI has evolved substantially over the past four decades. Using discharge International Classification of Diseases, Ninth Revision Clinical Modification (ICD-9-CM) codes, the National Hospital Discharge Survey (1979-2005) reported that age-adjusted AMI hospitalization rates increased from 215 cases per 100,000 persons in 1979-1981 to a peak of 342 in 1985-1987 before declining to 242 in 2003-2005 (2). Men and women displayed similar trends, though the AMI burden among men was nearly twice that of women. The Atherosclerosis Risk in Communities study, one of the largest and most rigorously validated population-based surveillance studies of AMI incidence in the United States, reported relatively stable rates of hospitalized AMI from 1987 to 1996 despite substantial declines in coronary heart disease mortality, suggesting that improved post-AMI survival contributed to this apparent paradox (3). Declines in AMI incidence then accelerated through 2008 despite adoption of conventional cardiac troponin assays that would have otherwise masked these improvements (3). Population-level data from Northern California (1999-2008) further illustrated this evolving landscape: ST-segment elevation myocardial infarction (STEMI) incidence declined 62% over the decade, whereas non–STEMI (NSTEMI) incidence initially rose, attributed to adoption of conventional cardiac troponin assays, before declining to near baseline levels once testing rates stabilized (4). An extended temporal analysis within the same healthcare system was conducted from 2008-2014 which demonstrated consistent declines in AMI rates across all subgroups including age, sex, and patients with diabetes (5). In contrast to the previous decade, the magnitude of the STEMI decline slowed, reaching a relative plateau by 2014.

The widespread transition to high-sensitivity cardiac troponin T and I (hs-cTn) assays following the U.S. Food and Drug Administration approval in 2017 has fundamentally altered the diagnostic and epidemiologic landscape of AMI. Defined by their ability to quantify troponin above the limit of detection in at least 50% of healthy individuals, hs-cTn assays offer substantially improved analytical sensitivity compared with conventional troponin assays, which themselves represented a major advance over creatine kinase-MB in both sensitivity and cardiac specificity (6). The enhanced sensitivity of hs-cTn enables detection of previously unrecognized cardiomyocyte injury. The Fourth Universal Definition of Myocardial Infarction (2018) formalized the distinction between myocardial injury—defined as any troponin elevation above the 99th percentile upper limit of a healthy reference population—and myocardial infarction, which requires evidence of a rise and/or fall in troponin in the setting of acute myocardial ischemia. The framework further classifies AMI into Type 1 (atherothrombotic) and Type 2 (supply-demand mismatch without acute atherothrombosis; T2MI), a distinction with important epidemiologic and therapeutic implications (7). Implementation of hs-cTn assays has increased the detection of AMI, particularly T2MI, as well as acute myocardial injury (8,9). By 2021, 32.6% of the 550 participating hospitals in the NCDR^®^ Chest Pain-MI Registry had adopted hs-cTn assays, an increase from 3.3% in 2019 (10). Given the rapid expansion of hs-cTn testing and strong guideline recommendations for its use in patients with suspected acute coronary syndrome (11), questions remain regarding the population-level implications of widespread utilization, particularly changes in testing volume and the recorded frequency and composition of AMI diagnoses. To our knowledge, no national-scale longitudinal EHR analysis has characterized documented AMI diagnosis rates and subtype composition during the concurrent expansion of hs-cTn testing and dedicated T2MI coding. Accordingly, the objectives of this study were to: 1). evaluate contemporary national trends in documented AMI diagnoses across the hs-cTn era; 2). characterize changes in AMI subtype composition, particularly the emergence of T2MI; 3). examine hs-cTn assay testing utilization and the proportion and composition of AMI diagnoses associated with hs-cTn-tested encounters.

## Methods

### Data Source

This retrospective serial cross-sectional study was performed using data from Epic Cosmos, a national multicenter electronic health record (EHR) database that integrates inpatient and outpatient data from participating U.S. healthcare organizations. As of July 2026, Cosmos included approximately 307 million patients, 21.6 billion encounters, and 2,197 hospitals (12). Epic reports that the database provides a broadly representative sample of patients who seek healthcare in the United States, based on comparisons with U.S. Census distributions across demographic and socioeconomic characteristics. Data were last queried in Epic Cosmos on July 27, 2026. This study used aggregate, de-identified data and did not involve access to individual patient-level information. Accordingly, the study did not constitute human subjects research and institutional review board approval and informed consent were not required. The data underlying this study were obtained from Epic Cosmos and are not publicly available from the authors.

### Identification and Characterization of Acute Myocardial Infarction

AMI diagnoses occurring between January 1, 2016, and December 31, 2024, among U.S. adults aged ≥18 years were identified from billed final diagnoses in Epic Cosmos. Any AMI was defined using ICD-10-CM category I21 and all descendant codes. Selected subtypes were defined as STEMI (I21.0*, I21.1*, I21.2*, and I21.3), NSTEMI (I21.4), and T2MI (I21.A1), consistent with the classification established by the Fourth Universal Definition of Myocardial Infarction (7). AMI subtypes were identified independently and were not required to be mutually exclusive. The dedicated I21.A1 code became available on October 1, 2017; therefore, T2MI was not separately identifiable in 2016 and was captured for only part of 2017.

The appropriate diagnosis position for identifying AMI varies by subtype. In a Taiwanese validation study, STEMI codes restricted to the first three diagnosis positions provided the best balance of sensitivity and positive predictive value (PPV), whereas NSTEMI and overall AMI— defined in that study as STEMI or NSTEMI—performed best when identified from any diagnosis position (13). Similarly, a validation study conducted within Kaiser Permanente Southern California found that principal-diagnosis AMI codes had a PPV of 86.8%, compared with 55.8% for nonprincipal codes; however, restricting ascertainment to the principal diagnosis excluded 14.8% of confirmed AMI events, defined as STEMI or NSTEMI (14). Because T2MI occurs secondary to an underlying precipitating condition and therefore may not be listed as the principal diagnosis, AMI was identified from billed final diagnoses in any position. This approach was applied consistently across all AMI subtypes.

### AMI Diagnosis Rates

Annual AMI diagnosis rates were estimated using the Patients model, where each patient contributed no more than once per calendar year for each outcome. The numerator comprised adult patients with a qualifying billed final AMI diagnosis during that year, and the denominator comprised all adults with at least one recorded encounter in Epic Cosmos during the corresponding year. Six strata were defined by age at encounter (18-44, 45-64, and ≥65 years) and recorded sex (male or female). Rates were directly standardized to the pooled age and sex distribution of the study population across 2016-2024 by multiplying each stratum-specific annual rate by its corresponding reference population weight and summing the weighted rates. The six reference rates were 0.174 for males aged 18-44 years, 0.231 for females aged 18-44 years, 0.145 for males aged 45-64 years, 0.187 for females aged 45-64 years, 0.114 for males aged ≥65 years, and 0.149 for females aged ≥65 years. Standardized rates were expressed per 100,000 patients. To characterize changes in the relative distribution of selected AMI subtypes, the age- and sex-standardized rate for each subtype was divided by the sum of the standardized NSTEMI, STEMI, and T2MI rates for the corresponding year. Because subtype diagnoses were ascertained independently and could overlap, these proportions represent the relative distribution of selected subtype rates rather than mutually exclusive proportions of patients with AMI. Patients with missing age or sex were excluded from the analyses.

### Identification of High-Sensitivity Cardiac Troponin Testing

Analyses of hs-cTn testing were conducted from January 1, 2017, through December 31, 2024, using the Encounters model. The first hs-cTn assay was cleared for clinical use in the United States in January 2017; therefore, 2017 was considered an initial adoption year. Qualifying encounters contained at least one recorded hs-cTn measurement, identified using LOINC codes 89579-7 for hs-cTn I or 67151-1 hs-cTn T, irrespective of the measured value. Multiple measurements within the same encounter were counted once, although individual patients could contribute more than one encounter. For each year, the proportion of hs-cTn-tested encounters associated with a documented AMI was calculated by dividing the number containing a billed final AMI diagnosis by the total number of qualifying hs-cTn-tested encounters. AMI diagnoses and hs-cTn testing were required to occur within the same encounter.

## Results

### Study Population

Across 2016-2024, the pooled study denominator included 821,859,867 patient-years. Of these, 43.3% were contributed by males and 56.7% by females; 40.5% were among adults aged 18-44 years, 33.2% among those aged 45-64 years, and 26.3% among those aged ≥65 years.

### Temporal Trends in Acute Myocardial Infarction Diagnosis Rates and Subtype Distribution

Age- and sex-standardized rates of documented AMI diagnoses increased from 343.1 per 100,000 adults in 2016 to 602.7 per 100,000 in 2024, representing a 75.7% increase. Rates initially increased through 2019, stabilized from 2019-2021, and subsequently rose further through 2024 (Figure 1). Trends differed substantially by AMI subtype. The standardized NSTEMI rate increased from 241.3 to 274.6 per 100,000 between 2016 and 2024, a 13.8% increase, although rates fluctuated over the study period and remained below the 2017 peak until 2023. The STEMI rate remained comparatively stable, decreasing from 111.9 to 107.2 per 100,000 (4.1% decrease). In contrast, the T2MI rate increased from 17.6 per 100,000 during its first year of partial capture in 2017 to 233.4 per 100,000 in 2024. From 2018, the first complete calendar year following adoption of the I21.A1 code, through 2024, the T2MI rate increased from 99.9 to 233.4 per 100,000, a 133.8% increase. By 2024, the total T2MI rate approached that of NSTEMI and was more than twice the STEMI rate.

**Figure 1.**
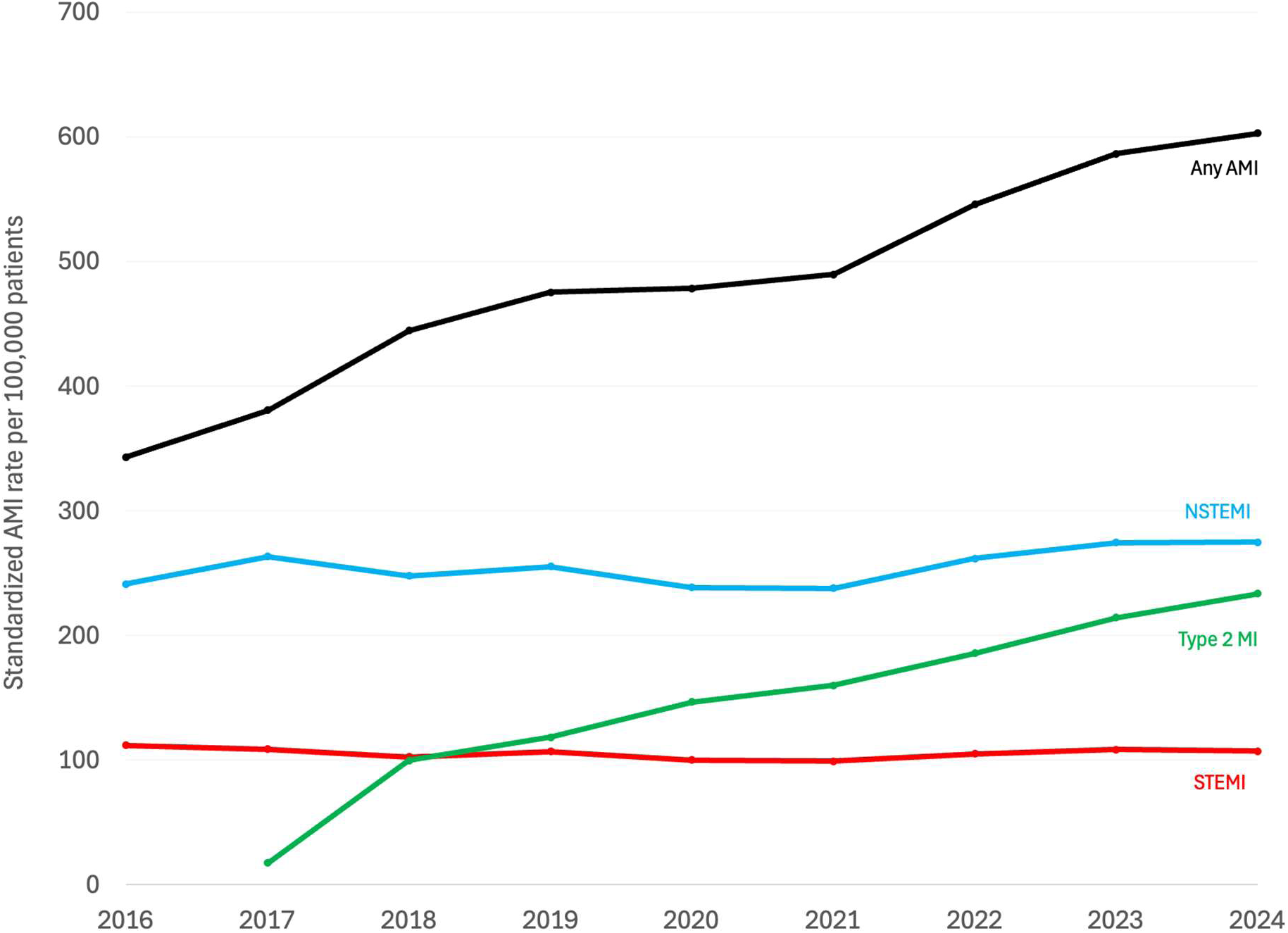
Age- and sex-standardized rates of acute myocardial infarction diagnoses, 2016-2024. Rates are shown for any AMI, NSTEMI, STEMI, and T2MI among adult patients. Rates are reported per 100,000 patients. The ICD-10-CM code for T2MI became effective in October 2017.

The relative proportion of AMI subtype rates changed substantially (Figure 2). In 2016, before T2MI was separately codable, NSTEMI and STEMI accounted for 68.3% and 31.7% of the summed standardized subtype rates, respectively. By 2024, these proportions had declined to 44.6% for NSTEMI and 17.4% for STEMI. T2MI accounted for 22.2% of the summed standardized subtype AMI diagnoses in 2018, the first complete year after introduction of the I21.A1 code, and increased to 37.9% by 2024.

**Figure 2.**
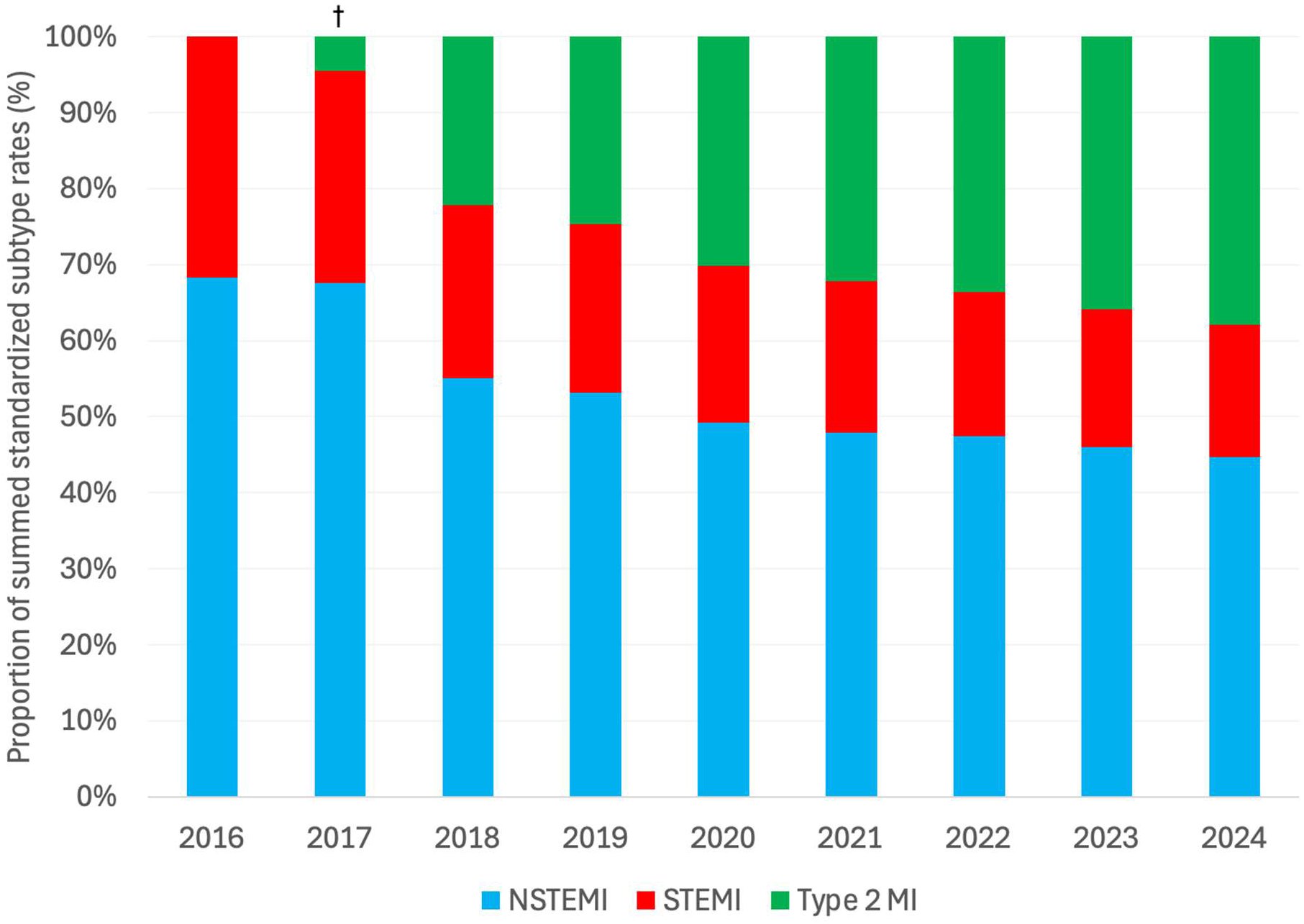
Relative distribution of age- and sex-standardized rates for selected acute myocardial infarction subtypes, 2016–2024. For each year, standardized NSTEMI, STEMI, and T2MI rates were expressed as percentages of their summed standardized rates. ^†^ICD-10-CM code I21.A1 for T2MI was introduced on October 1, 2017; therefore, T2MI was not separately identifiable in 2016 and was captured for only part of 2017.

### Age- and Sex-Stratified Acute Myocardial Infarction Diagnosis Rates

AMI diagnosis rates increased across all age and sex groups between 2016 and 2024, with higher rates among older adults and males throughout the study period (Figure 3). Among adults aged 18-44 years, rates increased from 87 to 142 per 100,000 among males and 27 to 57 per 100,000 among females.

**Figure 3.**
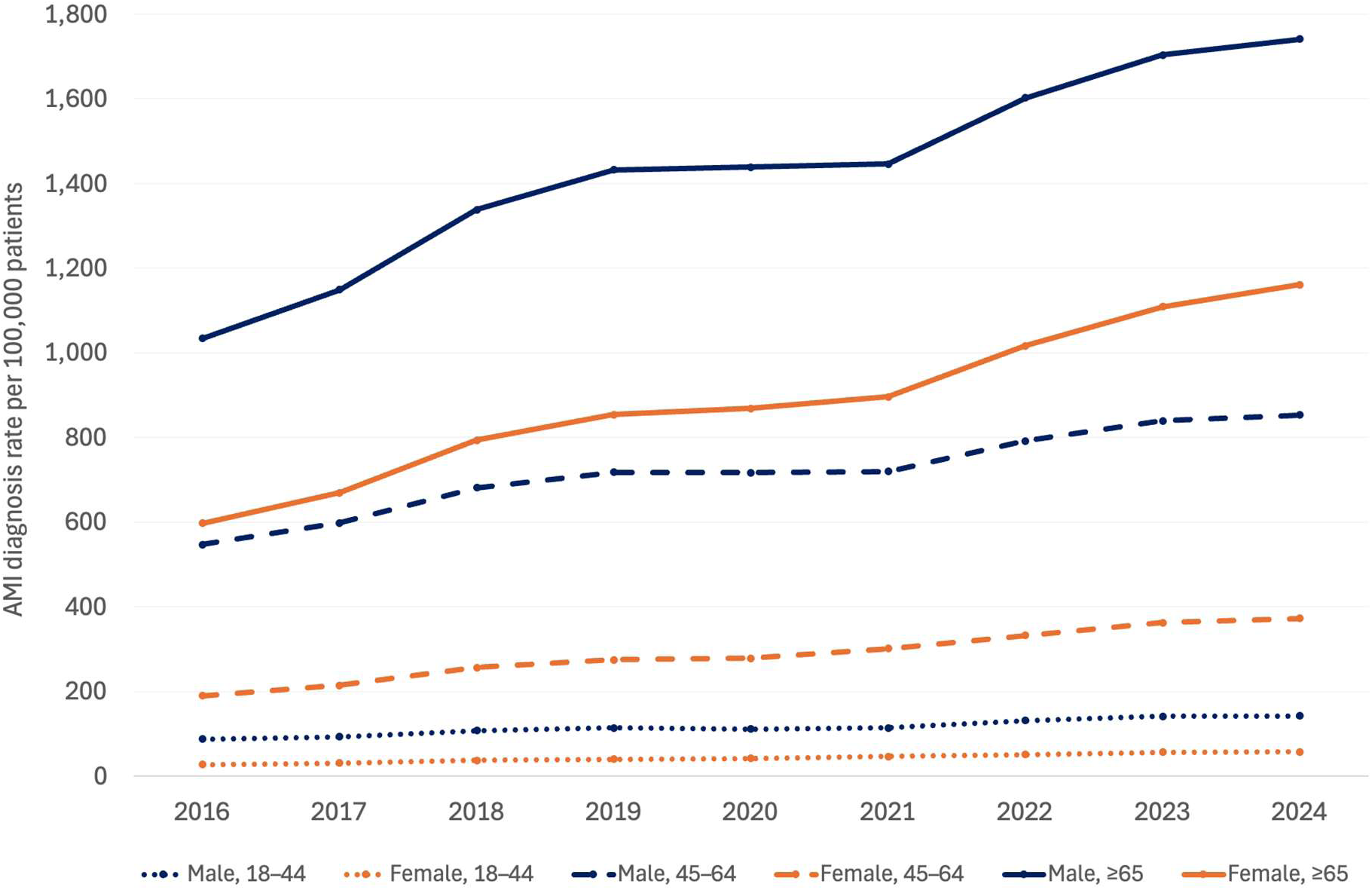
Age- and sex-specific acute myocardial infarction diagnosis rates, 2016–2024. Annual rates were calculated within each age- and sex-specific stratum and expressed per 100,000 adult patients. Any AMI was identified using ICD-10-CM code I21 and all descendant codes, including T2MI following the introduction of I21.A1 in October 2017.

Corresponding rates increased from 547 to 853 and from 190 to 372 per 100,000 among males and females aged 45-64 years, respectively, and from 1,034 to 1,741 and from 597 to 1,161 among those aged ≥65 years. Although absolute rates remained higher among males, relative increases were greater among females in every age group. From 2016-2024, relative rates increased by 63.2%, 55.9%, and 68.4% among males aged 18-44, 45-64, and ≥65 years, respectively, compared with 111.1%, 95.8%, and 94.5% among females. From 2016-2024, the male-to-female rate ratio narrowed from 3.22 to 2.49 among adults aged 18-44 years, from 2.88 to 2.29 among those aged 45-64 years, and from 1.73 to 1.50 among those aged ≥65 years.

### Acute Myocardial Infarction Diagnoses Among High-Sensitivity Cardiac Troponin-Tested Encounters, 2017-2024

Annual encounters involving at least one hs-cTn measurement increased from 187,949 in 2017 to 6,493,238 in 2024 (34.5-fold). The proportion of hs-cTn-tested encounters associated with any documented AMI diagnosis increased from 1.01% in 2017 to 4.74% in 2021 and remained stable thereafter (4.81% in 2024; Figure 4). All three subtype-specific proportions increased through 2021 but subsequently diverged: the proportions associated with NSTEMI and STEMI declined from 2.30% and 0.80% in 2021 to 1.99% and 0.63% in 2024 (relative declines of 13.5% and 21.1%), whereas the proportion associated with T2MI increased from 1.60% to 2.12% (relative increase of 32.3%), exceeding that of NSTEMI for the first time in 2024. Thus, from 2021 through 2024, hs-cTn-tested encounters increased 3.9-fold while the proportion associated with any AMI remained stable, as continued growth in T2MI was offset by declining proportions of NSTEMI and STEMI.

**Figure 4.**
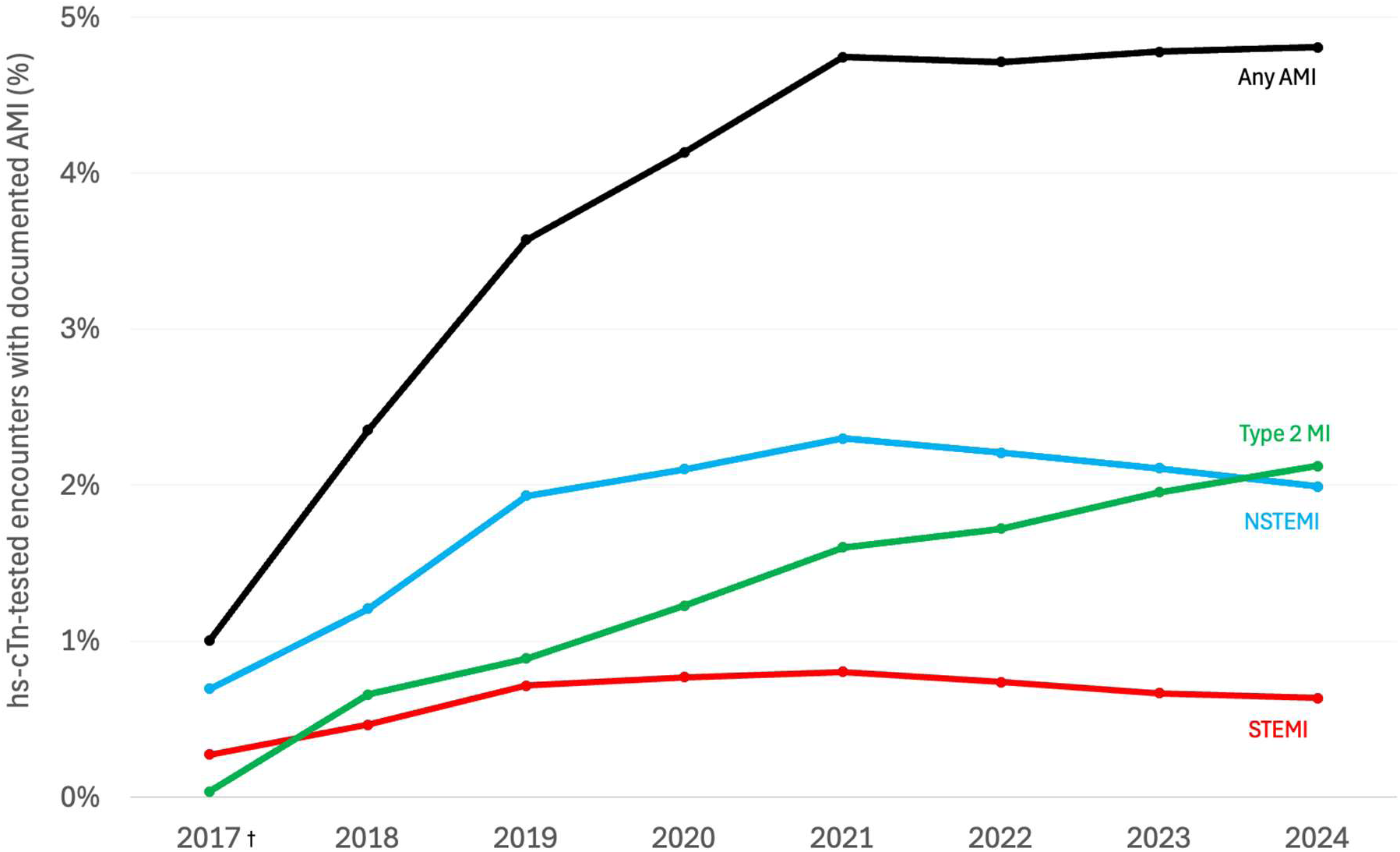
Acute myocardial infarction diagnoses among high-sensitivity cardiac troponin-tested encounters, 2017–2024. Annual proportions were calculated as the percentage of hs-cTn encounters associated with a documented diagnosis of any AMI, NSTEMI, STEMI, or T2MI during the same encounter. ^†^The first hs-cTn assay was cleared for clinical use in the United States in January 2017, and the dedicated ICD-10-CM code I21.A1 for T2MI became available in October 2017; therefore, 2017 represents an initial hs-cTn adoption year with partial-year ascertainment of T2MI.

## Discussion

### Principal Findings

In this national-scale EHR analysis, age- and sex-standardized rates of documented AMI diagnoses increased 75.7% from 2016 through 2024, driven predominantly by a 133.8% increase in documented T2MI beginning in 2018. In contrast, NSTEMI increased modestly and STEMI remained comparatively stable, resulting in a substantial shift in the recorded composition of AMI toward T2MI. Males consistently had higher absolute AMI diagnosis rates, although females experienced larger relative increases from lower baseline rates, and sex disparities narrowed over time. Concurrently, the annual number of encounters involving hs-cTn testing increased 34.5-fold from 2017 through 2024. The proportion of hs-cTn encounters associated with any AMI diagnosis initially increased but plateaued after 2021 despite continued growth in testing volume. During this plateau, the proportions associated with NSTEMI and STEMI declined, whereas the proportion associated with T2MI continued to increase.

### Findings in the Context of Prior Literature

Contemporary longitudinal EHR-based studies of AMI rates in the United States are limited, particularly during the era of widespread hs-cTn use and dedicated T2MI coding. Earlier studies from regional health systems, primarily spanning 2000 through 2008, demonstrated substantial declines in both NSTEMI and STEMI rates, attributed to improvements in cardiovascular risk-factor control (4). Follow-up analyses extending through 2014 found continued declines in NSTEMI across age, sex, and racial and ethnic groups, whereas declines in STEMI had begun to plateau (5). Another EHR-based study similarly reported declining AMI rates with a plateau among females from 2010 through 2014 (15).

In contrast to these earlier reports, documented AMI rates increased 75.7% in our study. This divergence was driven predominantly by a 133.8% increase in T2MI from 2018-2024, whereas NSTEMI increased by 13.8% and STEMI decreased by 4.1% across the full study period. Both NSTEMI and STEMI declined during the early COVID-19 period before subsequently rebounding, consistent with prior studies demonstrating reductions in AMI hospitalizations early in the pandemic (16,17).

These divergent trends extended to age- and sex-stratified rates. Previous studies have generally reported AMI rates approximately twice as high among males as females, with the sex disparity narrowing with increasing age and persisting across the study period (21). Our data corroborate the persistent male predominance and its attenuation with increasing age, but the male-to-female rate ratio narrowed over the study period within every age group, reflecting greater relative increases among females. This convergence occurred contemporaneously with the shift in diagnostic composition toward T2MI, although subtype-specific demographic trends were not examined in the present study.

Prior hs-cTn implementation studies support a disproportionate effect on the identification of T2MI. A community-based Wisconsin study found that implementation of a fifth-generation hs-cTn T assay from 2018-2019 more than doubled AMI diagnoses relative to the preceding fourth-generation assay, driven primarily by a 3.25-fold increase in T2MI. T1MI diagnoses increased by 70.6%, a less pronounced increase that was restricted to women (8). Similarly, implementation of the hs-cTn I assay in the High-STEACS randomized controlled trial reclassified 17% of patients with myocardial injury or infarction not identified by the contemporary assay. A prespecified secondary analysis demonstrated increases in T1MI of 11%, T2MI of 22%, and acute myocardial injury of 36%, without corresponding improvements in treatment or 1-year clinical outcomes (18,9).

Our findings extend these local implementation studies by characterizing longitudinal diagnostic trends across a large national EHR population during the concurrent expansion of hs-cTn testing and dedicated T2MI coding. Encounters involving hs-cTn testing expanded markedly from 2017-2024. The proportion of hs-cTn-tested encounters associated with any AMI diagnosis stabilized after 2021, yet the diagnostic composition continued to shift, with T2MI surpassing NSTEMI as the AMI subtype most frequently diagnosed among hs-cTn-tested encounters by 2024. The substantial growth in T2MI diagnoses likely reflects multiple overlapping processes, including enhanced detection of myocardial injury through hs-cTn testing, growing clinician familiarity with T2MI, and broader use of the dedicated I21.A1 code. Accordingly, the observed increase cannot solely be interpreted as increased incidence of T2MI.

### Interpretation and Clinical Implications

The trajectory of hs-cTn testing parallels prior experience with increasingly sensitive diagnostic strategies. Widespread adoption of computed tomography pulmonary angiography (CTPA) was followed by an 81% increase in the incidence of diagnosed pulmonary embolism with only minimal change in population mortality, alongside a 71% increase in complications of anticoagulation (19). However, this analogy is incomplete. Unlike potentially clinically inconsequential pulmonary emboli detected by CTPA, hs-cTn elevation outside T1MI is not necessarily a false-positive finding and carries prognostic information across multiple cardiovascular and systemic conditions (20). Furthermore, hs-cTn implementation in one community-based study was associated with only a modest increase in coronary angiography, without corresponding increases in hospital admission, echocardiography, or stress testing (8). The principal consequence may therefore be less procedural than diagnostic and therapeutic: hs-cTn testing increases recognition of myocardial injury and T2MI—conditions associated with substantial long-term mortality—but their management remains largely unsupported by randomized trial evidence (21).

These findings underscore the need for consistent implementation and trial-based evaluation of consensus-based diagnostic and management pathways that distinguish T1MI from T2MI and guide subsequent cardiovascular evaluation. As hs-cTn utilization continues to expand and the documented composition of AMI shifts toward T2MI, randomized trials are needed to determine whether structured investigative strategies and mechanism-directed treatments improve outcomes in this heterogeneous population.

### Strengths

This study used a large, national-scale, multicenter EHR population spanning the United States and extended earlier regional EHR investigations into the contemporary era of dedicated T2MI coding and widespread hs-cTn adoption. AMI ascertainment was methodologically aligned with prior EHR studies to facilitate historical comparison. Direct age- and sex-standardization facilitated improved temporal demographic comparability. Separate characterization of any AMI, NSTEMI, STEMI, and T2MI allowed subtype-specific trends to emerge, while age- and sex-stratified analyses included younger adults who have been less extensively characterized in prior studies. Finally, the encounter-level analysis uniquely characterized the substantial expansion of hs-cTn testing and the subsequent shift in documented AMI diagnoses toward T2MI.

### Limitations

Several limitations should be considered. First, AMI diagnoses were identified using billed ICD-10-CM codes without clinical adjudication against the Universal Definition of Myocardial Infarction. Prior validation studies have demonstrated variable performance across individual codes and diagnosis positions. Billed final diagnoses in any coding position were deliberately chosen to capture T2MI occurring secondary to an underlying precipitating condition; however, this strategy may have lower predictive value than restricting AMI diagnoses to the principal discharge diagnosis position. Because AMI subtypes were independently ascertained and were not required to be mutually exclusive, the total summed subtype rates modestly exceeded the any-AMI rate, potentially indicating dual or duplicate subtype coding. Additionally, the serial cross-sectional design could not distinguish incident from recurrent AMI; therefore, the reported rates represent annual documented AMI diagnoses rather than AMI incidence. Given that Epic Cosmos includes care-seeking patients from participating healthcare organizations rather than a probability sample of the U.S. population, changes in participating organizations, patient composition, and healthcare utilization may have influenced both numerators and denominators. Because Cosmos data were collected for clinical care rather than specifically for this research question, incomplete or missing EHR data and unmeasured temporal differences in patient and healthcare-system characteristics may have introduced residual bias. The direction and magnitude of these potential biases could not be quantified from available aggregate data. Finally, hs-cTn testing was identified through LOINC mappings rather than independently verified. Testing indications, measured concentrations, and serial changes were unavailable. Accordingly, the parallel expansion of hs-cTn testing and documented T2MI remains descriptive and cannot establish causality.

### Conclusion

In this national-scale Epic Cosmos EHR analysis, documented acute myocardial infarction rates increased substantially from 2016 through 2024, driven predominantly by growth in Type 2 myocardial infarction, while NSTEMI increased modestly and STEMI declined. Concurrently, encounters involving high-sensitivity cardiac troponin increased 34.5-fold from 2017 through 2024, and T2MI surpassed NSTEMI as the most frequently diagnosed subtype among tested encounters by 2024, even as the proportion of tested encounters associated with any acute myocardial infarction remained relatively stable after 2021. These findings characterize an evolving diagnostic composition of acute myocardial infarction toward Type 2 myocardial infarction and underscore the need for evidence-based approaches to its evaluation and management.

## Data Availability

The data underlying this study were obtained from Epic Cosmos and are not publicly available from the authors.

## Acknowledgments

None

## Non-standard Abbreviations and Acronyms

AMI: Acute Myocardial Infarction
CTPA: Computed Tomography Pulmonary Angiography
EHR: Electronic Health Record
ICD-9-CM: International Classification of Diseases, Ninth Revision, Clinical Modification
ICD-10-CM: International Classification of Diseases, Tenth Revision, Clinical Modification
NSTEMI: Non–ST-segment elevation myocardial infarction
PPV: Positive Predictive Value
STEMI: ST-segment elevation myocardial infarction
T1MI: Type 1 Myocardial Infarction
T2MI: Type 2 Myocardial Infarction

